# Development and validation of an XGBoost model with SHAP-based interpretability and a web-based calculator for predicting extrauterine growth restriction in preterm infants

**DOI:** 10.64898/2026.04.01.26349838

**Authors:** Zheng Xu, Chun-Lan Yu, Jing-Xia Zhang

**Affiliations:** College of Mathematics, Beijing Normal University, Beijing, China; Neonatal Intensive Care Unit, Obstetrics and Gynecology Hospital, Fudan University, Shanghai, China

**Keywords:** Extrauterine growth restriction, Preterm infants, Machine learning, XGBoost, SHAP, Risk prediction model, Web-based calculator

## Abstract

**Background:** Extrauterine growth restriction (EUGR) is a common and clinically significant complication among preterm infants, contributing to adverse neurodevelopmental and metabolic outcomes. Early and individualized risk prediction remains challenging. This study aimed to develop and validate an interpretable machine learning model for early prediction of EUGR using routinely available clinical variables, and to implement a user-friendly web-based calculator for clinical use.

**Methods:** We retrospectively analyzed 1,431 preterm infants admitted within 24 hours after birth to our hospital between May 2020 and March 2025. Infants from the Yangpu campus (n=863) formed the training set, and those from the Huangpu campus (n=568) formed the validation set. Early clinical variables available within 48–72 hours were screened using the Boruta algorithm. Logistic regression, XGBoost, random forest, decision tree, and support vector machine models were developed and compared. Model performance was evaluated using area under the curve (AUC), accuracy, sensitivity, specificity, F1 score, and Brier score. SHapley Additive exPlanations (SHAP) were applied to assess global and individual feature contributions, nonlinear effects, and interactions. A web-based calculator was constructed based on the optimal model.

**Results:** Nine variables were identified as important predictors: birth weight, small for gestational age status, gestational age, breastfeeding, multiple gestation, neonatal respiratory distress syndrome, patent ductus arteriosus, maternal hypertension, and maternal group B Streptococcus infection. Among the five models, XGBoost achieved the best performance in the validation set (AUC 0.922, accuracy 0.849, Brier score 0.108). SHAP analysis showed that low birth weight, small for gestational age, maternal group B Streptococcus infection, and patent ductus arteriosus were major risk factors, while breastfeeding was protective. Notable nonlinear and interactive effects were observed, particularly between birth weight and gestational age and between breastfeeding and patent ductus arteriosus. The web-based calculator provides real-time individualized risk estimation and visualized interpretation.

**Conclusions:** An interpretable XGBoost-based model and web calculator were successfully developed and validated for early prediction of EUGR in preterm infants. This tool may support clinicians in identifying high-risk infants and guiding individualized nutritional and clinical management.

## Introduction

Postnatal nutritional status in preterm infants, particularly extrauterine growth restriction (EUGR), remains a significant focus and challenge in neonatal research [1]. Studies have demonstrated that EUGR adversely affects not only short-term physical growth and complications in preterm infants but also their long-term health outcomes. It is associated with an increased risk of impaired neurocognitive development during childhood, as well as a heightened predisposition to metabolic syndromes, including obesity, diabetes, and hypertension, later in childhood and even into adulthood [2,3]. Therefore, the early and accurate identification of preterm infants at high risk for EUGR and the timely implementation of interventions represent a research priority with critical implications for both their immediate and long-term health.

In recent years, numerous studies have been dedicated to identifying risk factors for EUGR in preterm infants, leading to the recognition of several factors significantly associated with an increased risk of EUGR. However, many studies rely on univariate screening followed by multivariable logistic regression to build prediction models [4–6]. We previously conducted a similar study [7] (see Supplementary File 1 for the full text of this Chinese-language article), and developed a nomogram based on our institutional data, but its performance in practice was suboptimal. While various factors may influence real-world application, the type and performance of the constructed model are among the primary reasons. Nomograms based on logistic regression assume a linear relationship between independent variables and the log-odds of the outcome. Consequently, they often exhibit poor fit when actual nonlinear associations or interaction effects exist among risk factors within the model [8]. Existing reported EUGR prediction models typically estimate the probability of EUGR occurrence under specific variable conditions but fail to explain the individual contribution of each risk factor to the prediction result at the individual subject level [9, 10].

With the progressive integration of artificial intelligence into the medical field, machine learning algorithms have often demonstrated superior performance in health assessment studies involving complex, high-dimensional clinical data with intricate variable relationships [11–14]. Given the limited practical utility and room for performance improvement in our previous model, this study adopts a refined methodology. We leverage interpretable machine learning techniques to construct an Extreme Gradient Boosting (XGBoost) model capable of capturing nonlinear relationships among feature variables for EUGR prediction. Furthermore, we employ SHapley Additive exPlanations (SHAP) values for interpretability analysis, including quantifying the contribution of individual features, visualizing the strength of risk factors in predictions for single samples, and exploring nonlinear and interactive effects among features. Finally, we develop a web-based, real-time EUGR risk calculator centered around the backend XGBoost prediction model to facilitate clinical application. This work aims to provide healthcare professionals with a scientific basis for early, personalized clinical management decisions, ultimately improving growth and developmental outcomes for preterm infants.

## Objects and methods

### Study population

This study used data that were largely aligned with our previous work, with additional cases included from the Huangpu campus [7]. Preterm infants born and hospitalized at the Yangpu campus of our hospital between May 2020 and March 2025 were retrospectively selected as the training set for model development. Preterm infants born and hospitalized at the Huangpu campus between May 2022 and March 2025 served as the validation set for assessing the model’s predictive performance.

Inclusion criteria were: 1) hospital admission within 24 hours of birth; 2) gestational age between 28 weeks and 36^+6^ weeks; 3) length of hospital stay at least 4 days; and 4) availability of complete clinical data. Exclusion criteria were: 1) in vitro fertilization conception; 2) presence of congenital genetic diseases; 3) death during hospitalization or discharge against medical advice; and 4) maternal history of major organic diseases or severe smoking/alcohol use.

### Sample size estimation and data collection

Sample size estimation and data collection were performed according to the methodology described in our previous work [7]. Briefly, the minimum required sample size for the training set was 354, and for the validation set, it was 152. All included variables were obtainable early after birth (within 48~72 hours). These included: maternal age at pregnancy, delivery via cesarean section, multiple gestation, corticosteroid use, and maternal comorbidities during pregnancy including diabetes, hypertension, thyroid disease, and Group B Streptococcus (GBS) infection. Preterm infant clinical data included: gestational age, birth weight, sex, 1-minute Apgar score, small for gestational age (SGA) status, early breastfeeding, neonatal respiratory distress syndrome (NRDS), neonatal asphyxia, patent ductus arteriosus (PDA), hyperbilirubinemia, and anemia.

### Statistical methods

#### Descriptive statistics

Data analysis was performed using STATA 18.0 statistical software. Categorical data are presented as frequencies and percentages, and group comparisons were conducted using the χ^2^ test. Continuous variables with non-normal distributions are presented as medians with interquartile ranges, and group comparisons were performed using the Mann–Whitney U test. A small proportion of missing data was identified in the dataset. To address this, we performed multiple imputation using the mice package in R software, which employs a random forest-based approach. Five imputed datasets were generated and pooled using Rubin’s rules for subsequent analyses. A *P*-value < 0.05 was considered statistically significant.

#### Model development and evaluation

Using R software (version 4.5.0) and the Boruta package, variable screening was performed via the Boruta algorithm. Features confirmed as important by the Boruta algorithm were retained for model building. Logistic regression served as the baseline model. Four nonlinear machine learning methods—XGBoost, random forest, decision tree, and support vector machine (SVM)—were used to construct and train the prediction models, and their performance was compared with that of the baseline model. Model performance was evaluated using metrics including accuracy, sensitivity, specificity, F1 score, area under the receiver operating characteristic (ROC) curve (AUC), and the Brier score.

#### Model interpretability analysis

Interpretability analysis for the XGBoost model was performed using SHAP. SHAP values were calculated using the shapviz package in R 4.5.0. A global feature importance bar plot was generated using ggplot2 to visualize the overall contribution of each variable to EUGR prediction. A beeswarm plot was used to visualize the distribution of SHAP values for each feature. Individual waterfall plots were created to explain the direction and magnitude of each feature’s contribution to the prediction outcome for representative individual samples. SHAP dependence plots for individual features were drawn to explore potential nonlinear effects and interaction effects between variables.

#### Development of a web-based real-time EUGR risk calculator

To facilitate clinical application, an interactive web application prototype was developed using Python 3.11 and the Streamlit 1.28 framework. This calculator uses the backend XGBoost prediction model as its core. It features a user-friendly front-end interface that accepts user inputs for the early clinical features selected by the Boruta algorithm, computes the EUGR risk probability in real-time, and returns the result. The system can be deployed on a server and is accessible via a specified URL on multiple platform browsers.

#### Model calibration and evaluation of clinical net benefit

To further assess the clinical utility of the prediction model, calibration curves were used to evaluate model calibration. Decision curve analysis (DCA) was employed to quantify the net benefit of the XGBoost model across different decision thresholds. The decision curve for the XGBoost model on the validation set was calculated using the rmda package in R 4.5.0 software, assessing the model’s practical value in clinical decision-making.

## Results

### General characteristics of the study cohort

A total of 1,431 preterm infants from both campuses were included in this study, of which 488 developed EUGR, yielding an incidence rate of 34.1%. The baseline characteristics of the infants in the two groups are presented in Table 1. Univariate analysis indicated that 13 indicators showed statistically significant differences (*P* < 0.05). After grouping by campus, the Yangpu campus cohort formed the training set, comprising 863 cases (299 with EUGR), while the Huangpu campus cohort formed the validation set, comprising 568 cases (189 with EUGR). The sample sizes for both sets significantly exceeded the calculated minimum required sample sizes, thus meeting the study requirements. No statistically significant difference was observed in the incidence of EUGR between the training and validation sets (*P* = 0.592).

**Table 1.** Baseline characteristics of the study participants[M(P_25_, P_75_) or *n*(%)]

| Factors | Total(n=1431) | EUGR(n=488) | Non-EUGR(n=943) | $\chi^2/Z$ | P | Training set(n=863) | Validation set(n=568) |
| --- | --- | --- | --- | --- | --- | --- | --- |
| Birth weight | 2022(1700,2270) | 1612(1367,1870) | 2204(1990,2440) | 23.843 | <0.001 | 1998(1700,2270) | 2008(1700,2270) |
| Gestational age | 33.6(32.5,35.2) | 32.9(31.4,34.5) | 34.0(33.1,35.4) | 9.051 | <0.001 | 33.6(32.4,35.2) | 33.6(32.5,35.2) |
| Multiple gestation | 410(28.65%) | 156(31.97%) | 254(26.94%) | 3.983 | 0.046 | 270(31.29%) | 140(24.65%) |
| Male sex | 792(55.35%) | 243(49.80%) | 549(58.22%) | 9.232 | 0.002 | 478(55.39%) | 314(55.28%) |
| Apgar 1 min | 107(7.48%) | 45(9.22%) | 62(6.57%) | 3.256 | 0.071 | 67(7.76%) | 40(7.04%) |
| SGA | 355(24.81%) | 267(54.71%) | 88(9.33%) | 355.044 | <0.001 | 209(24.22%) | 146(25.70%) |
| NRDS | 438(30.61%) | 179(36.68%) | 259(27.47%) | 12.856 | <0.001 | 295(34.18%) | 143(25.18%) |
| Anemia | 243(16.98%) | 115(23.56%) | 128(13.57%) | 22.774 | <0.001 | 147(17.03%) | 96(16.90%) |
| Breastfeeding | 588(41.09%) | 127(26.02%) | 461(48.88%) | 69.437 | <0.001 | 366(42.41%) | 222(39.08%) |
| Neonatal asphyxia | 127(8.87%) | 44(9.02%) | 83(8.80%) | 0.018 | 0.892 | 70(8.11%) | 57(10.04%) |
| Hyperbilirubinemia | 1261(88.12%) | 435(89.14%) | 826(87.59%) | 0.735 | 0.391 | 765(88.64%) | 496(87.32%) |
| PDA | 293(20.48%) | 167(34.22%) | 126(13.36%) | 85.936 | <0.001 | 185(21.44%) | 108(19.01%) |
| Maternal age | 32.0(28.0,36.0) | 32.0(28.0,36.0) | 32.0(28.0,36.0) | -0.217 | 0.828 | 32.0(28.0,36.0) | 32.1(28.0,36.0) |
| GBS infection | 316(22.08%) | 182(37.30%) | 134(14.20%) | 99.603 | <0.001 | 205(23.75%) | 111(19.54%) |
| Maternal hypertension | 319(22.29%) | 161(32.99%) | 158(16.75%) | 48.941 | <0.001 | 180(20.86%) | 139(24.47%) |
| Maternal diabetes | 319(22.29%) | 113(23.16%) | 206(21.85%) | 0.319 | 0.572 | 193(22.36%) | 126(22.18%) |
| Maternal thyroid disease | 210(14.68%) | 99(20.29%) | 111(11.77%) | 18.625 | <0.001 | 105(12.17%) | 105(18.49%) |
| Corticosteroid use | 216(15.09%) | 59(12.09%) | 157(16.65%) | 5.215 | 0.022 | 128(14.83%) | 88(15.49%) |
| Cesarean delivery | 757(52.90%) | 248(50.82%) | 509(53.97%) | 1.286 | 0.257 | 493(57.13%) | 264(46.48%) |

### Feature selection in the training set

The Boruta algorithm was applied to screen all 19 potential influencing factors, as illustrated in Fig. 1. Features displayed in green were confirmed as important by the Boruta algorithm and were retained; those in red were deemed unimportant and could be removed; and those in yellow were of tentative importance. The final selection resulted in nine independent variables, shown in green, being retained for model construction: NRDS, multiple gestation, breastfeeding, maternal hypertension, PDA, maternal GBS infection, gestational age, SGA, and birth weight.

**Fig. 1.**
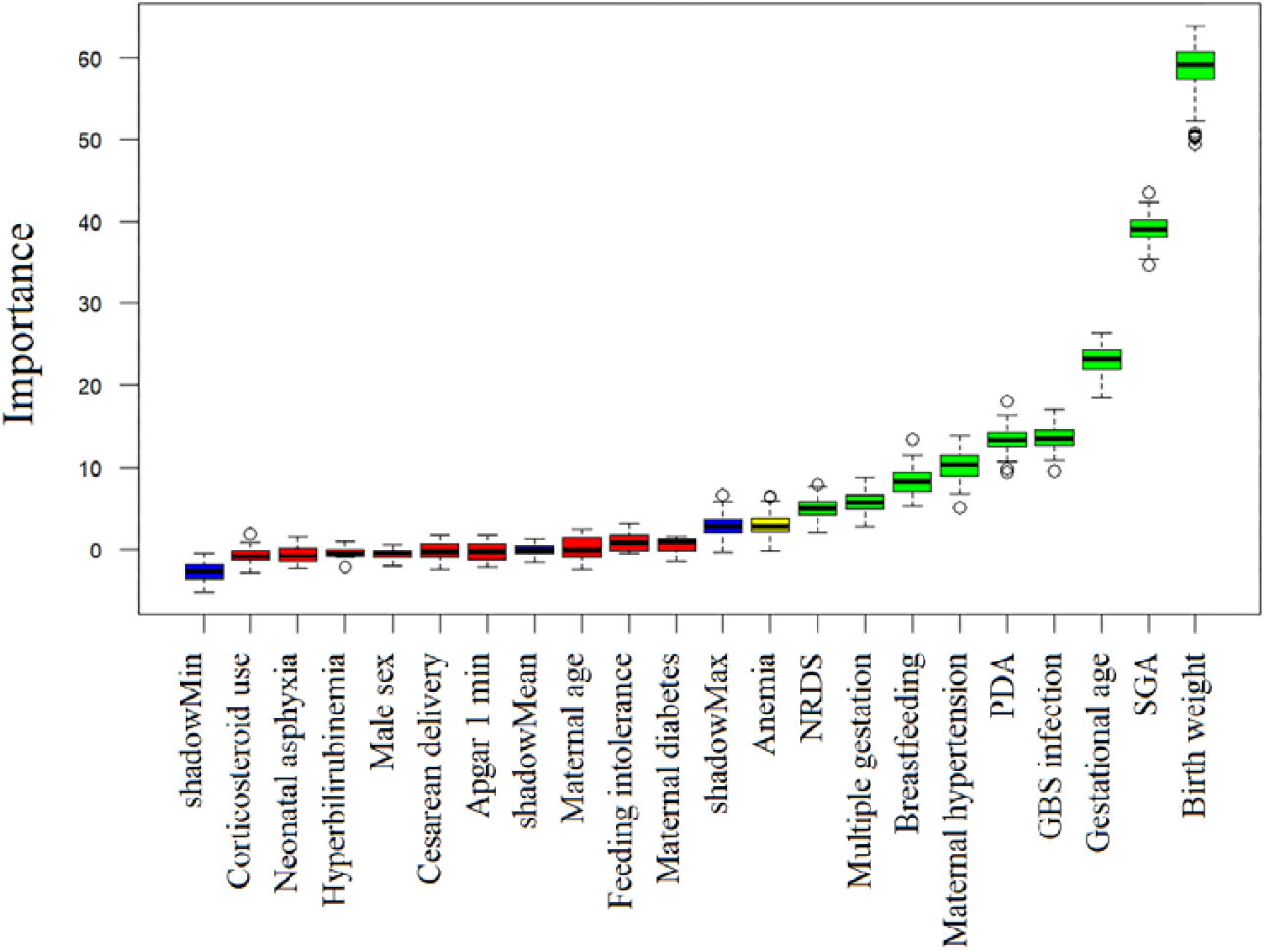
Boruta feature selection on the training set.

### Development and comparison of EUGR prediction models

Based on the nine features selected by the Boruta algorithm, five distinct models—Logistic Regression, XGBoost, Random Forest, Decision Tree, and SVM—were constructed on the training set. Their predictive performances were subsequently evaluated on the validation set, as detailed in Table 2. The results indicated that the XGBoost model demonstrated superior performance compared to the other four models. Graphical comparisons of model performance and ROC curves are provided in Supplementary Figs. S1 and S2.

**Table 2.** Comparison of the predictive performance of five machine learning models on the validation set.

| Model | Accuracy | Sensitivity | Specificity | F1 | AUC | Brier score |
| --- | --- | --- | --- | --- | --- | --- |
| XGBoost | 0.849 | 0.757 | 0.895 | 0.769 | 0.922 | 0.108 |
| Decision tree | 0.849 | 0.778 | 0.884 | 0.774 | 0.903 | 0.116 |
| Random forest | 0.842 | 0.773 | 0.876 | 0.764 | 0.900 | 0.122 |
| Logistic regression | 0.817 | 0.635 | 0.908 | 0.698 | 0.899 | 0.139 |
| SVM | 0.817 | 0.688 | 0.881 | 0.714 | 0.894 | 0.129 |

### Global and individual interpretability of the XGBoost model

To enhance the interpretability of the optimal XGBoost model, a feature importance bar plot and a beeswarm plot were generated. Fig. 2A lists the nine risk factors ranked by their mean absolute SHAP values, in descending order: birth weight, SGA, maternal GBS infection, PDA, gestational age, maternal hypertension, breastfeeding, multiple gestation, and NRDS. Fig. 2B illustrates the direction and magnitude of each feature’s contribution to the EUGR risk prediction. A positive SHAP value increases the risk, while a negative value decreases it. The plot shows that higher birth weight and greater gestational age are protective factors (negative SHAP values), whereas lower birth weight and smaller gestational age are high-risk factors (positive SHAP values). SGA, maternal GBS infection, PDA, maternal hypertension, and multiple gestation were all identified as risk factors for EUGR, while breastfeeding was a protective factor. The data points for NRDS are clustered near zero, suggesting a relatively weaker influence on EUGR. Individual waterfall plots were used to explain the contribution of each feature to the model’s prediction for single samples (Fig. 3). Fig. 3A represents a high-risk predicted case (ID2); Fig. 3B a low-risk predicted case (ID1); and Fig. 3C a borderline-risk predicted case (ID5). These plots respectively display the direction and size of each feature’s contribution for these individual predictions.

**Fig. 2.**
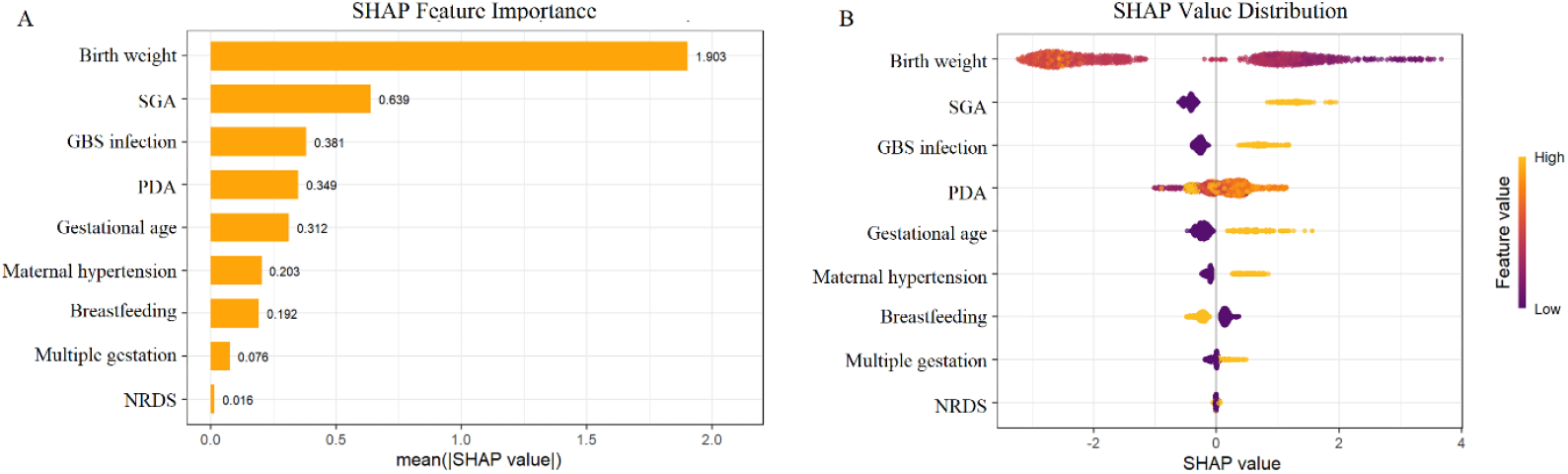
Global interpretability of the XGBoost model using SHAP. A: Feature importance bar plot; B: Beeswarm plot of feature contributions

**Fig. 3.**
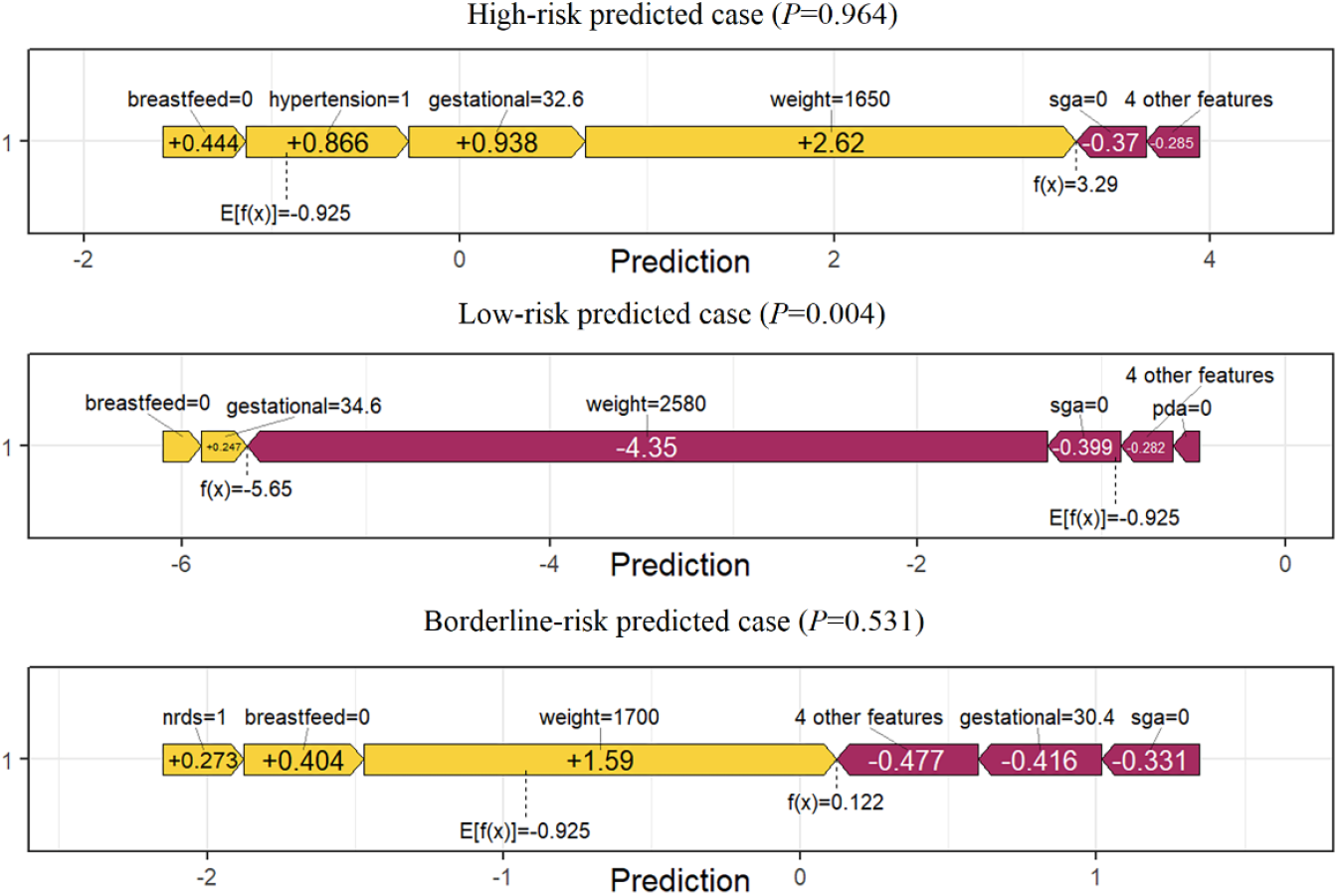
Waterfall plots for individual interpretation in the XGBoost model. A: High-risk predicted case (ID2); B: Low-risk predicted case (ID1); C: Borderline-risk predicted case (ID5)

### Preliminary exploration of nonlinear and interaction effects in EUGR prediction

The SHAP dependence plots derived from the XGBoost model (Fig. 4) revealed notable nonlinear effects among the continuous variables. Birth weight exhibited a pronounced nonlinear relationship: within the range of approximately 800g to 2000g, the SHAP value decreased rapidly from around 6 to 0 as birth weight increased. For birth weights above 2000g, the SHAP values plateaued around −4, indicating no further substantial decrease in risk with increasing weight (Fig. 4A). In contrast, gestational age demonstrated a weaker nonlinear effect in the SHAP dependence plot, with most data points scattered within the SHAP value range of −2 to 2 (Supplementary Fig. S3). Further assessment of interaction effects revealed a marked synergistic effect between birth weight and gestational age. Specifically, lower gestational age substantially increased the EUGR risk in infants with low birth weight, with SHAP values reaching approximately 6 (the deep-purple points cluster at higher positive SHAP values). Conversely, the data points for infants with both higher gestational age and higher birth weight largely overlapped, suggesting no additive protective effect. A significant interaction was also observed between breastfeeding and PDA (Fig. 4B). The protective effect of breastfeeding against EUGR was more pronounced in preterm infants with PDA compared to those without PDA (the yellow points are concentrated at more negative SHAP values). Conversely, the presence of PDA in infants who were not breastfed was associated with a notably higher probability of EUGR (the yellow points exhibit higher positive SHAP values). Interaction surface plots (Figs. 4D, 4E) visually demonstrate how the influence of one feature on risk changes according to the value of another feature. Other features also exhibited some degree of nonlinear associations and interaction effects, but these were comparatively weaker (Supplementary Figs. S3, S4).

**Fig. 4.**
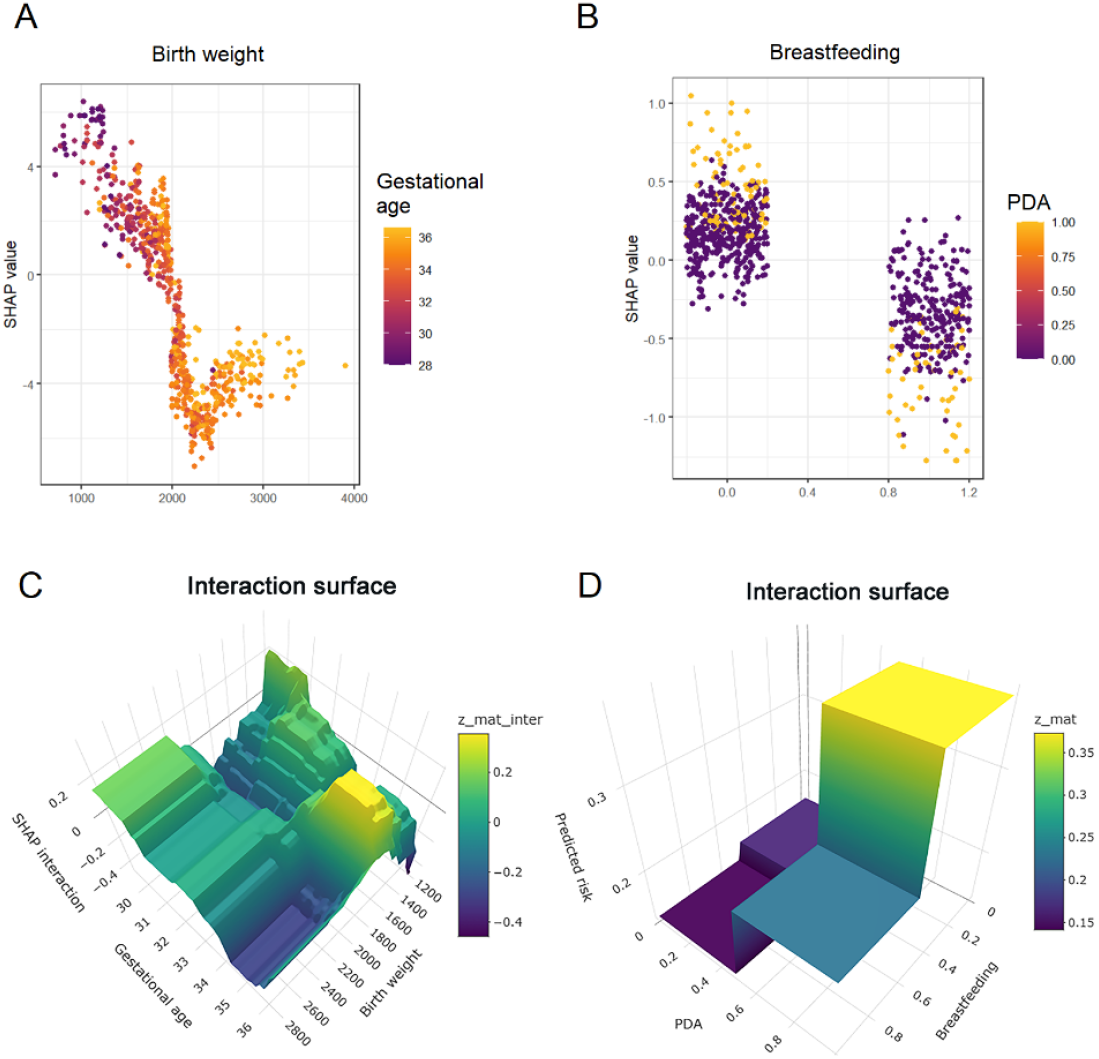
SHAP Dependence and Interaction Surface Plots. A: SHAP dependence plot for birth weight; B: SHAP dependence plot for breastfeeding; C: Interaction surface plot for birth weight and gestational age; D: Interaction surface plot for breastfeeding and PDA

### Development and validation of the web-based real-time EUGR risk calculator

This study successfully developed an interactive web application prototype, translating the EUGR risk prediction model into a potential clinical tool. The executable package is available as Supplementary File 2. As shown in Fig. 5A, the calculator provides an intuitive interface where clinicians can easily input key early clinical features using sliders, drop-down menus, and other form elements. The system then computes and displays the individual patient’s EUGR risk probability in real time, along with a breakdown of the specific risk factors contributing to that individual’s prediction. The calibration curve for the XGBoost model plotted on the validation set showed good agreement with the ideal curve (Fig. 5B). This assessment indicates that the model is well-calibrated, with predicted probabilities of EUGR closely matching the actual observed frequencies. The DCA curve (Fig. 5C) demonstrated that the model provides a higher net benefit across a wide range of high-risk thresholds (0.1 to 0.9) in the validation set, suggesting its potential value for clinical application.

**Fig. 5.**
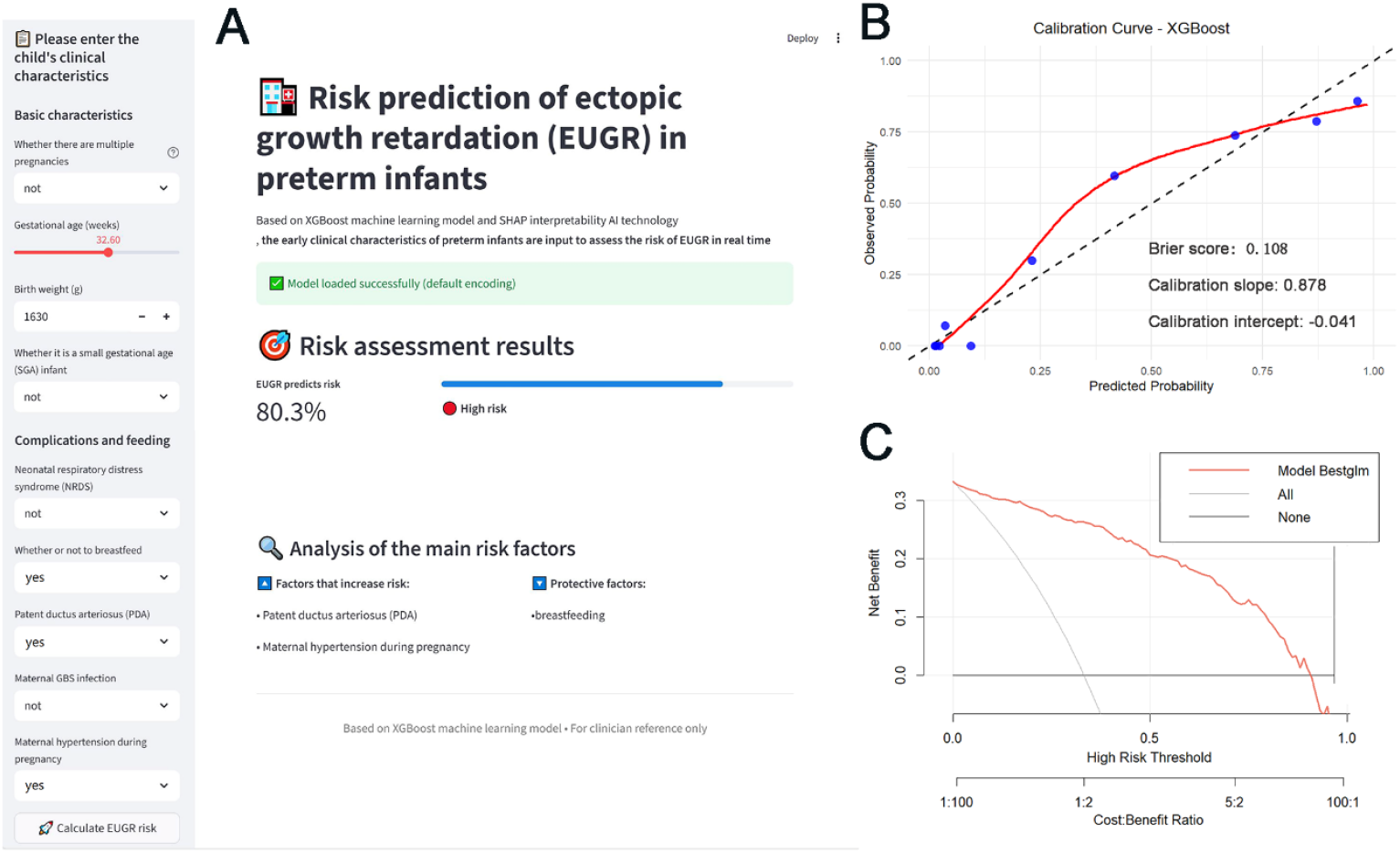
Clinical application and validation of the XGBoost-based EUGR risk prediction model. A: Interactive interface of the web-based real-time EUGR risk calculator; B: Calibration curve of the XGBoost model on the validation set; C: DCA of the XGBoost model on the validation set.

## Discussion

This study successfully developed and validated an interpretable machine learning model based on XGBoost for predicting the risk of EUGR in preterm infants, subsequently deploying it as a web-based calculator. The model demonstrated excellent predictive performance (AUC=0.922). Interpretability analysis using SHAP values quantified the contribution of individual risk factors to the occurrence of EUGR and also examined nonlinear and interaction effects among them, which are relatively uncommon in traditional studies of EUGR risk factors. This provides new insights for a deeper understanding of the roles various risk factors play in EUGR development.

In this study, the Boruta algorithm ultimately selected nine feature variables. Through global SHAP interpretation, the ranking and direction of each feature’s contribution to EUGR risk were clarified, offering a more refined basis for clinical risk assessment. Consistent with our previous work, birth weight, SGA, PDA, maternal hypertension, and maternal GBS infection were again confirmed as independent risk factors for EUGR, while breastfeeding demonstrated a consistent protective effect, the mechanisms of which have been detailed in our previously published literature [7]. Since Lasso regression is a generalized linear model, it struggles to capture nonlinear and interaction effects. The set of variables it selects is optimal for building a linear model but may omit many variables carrying important nonlinear signals [15, 16]. Therefore, this study employed the Boruta algorithm, which is based on random forests, to facilitate the identification of variables exhibiting any form of association, linear or nonlinear [17, 18]. The nine variables confirmed as important by Boruta also included gestational age, multiple gestation, and NRDS. Some scholars posit that smaller preterm infants have deficient energy reserves (such as fat and glycogen) at birth, are more susceptible to early respiratory and circulatory complications that restrict feeding, and have immature gut function, all of which can contribute to inadequate intake of key nutrients like protein and energy, leading to EUGR [19, 20]. Although gestational age was excluded by Lasso regression in the multifactorial linear model of our previous study, it was retained as an important feature in the current XGBoost model. Furthermore, our model identified multiple gestation and NRDS as independent risk factors for EUGR. Multiple gestation is often associated with competitive intrauterine growth restriction, a congenital disadvantage that may persist after birth, increasing the difficulty of postnatal catch-up growth and the risk of EUGR [21, 22]. NRDS, a common and severe respiratory condition in preterm infants, likely indirectly impedes normal physical growth through increased energy expenditure, metabolic stress, and potential disruption to the establishment of early enteral nutrition [23, 24]. These findings, which were not fully apparent in the previous linear model, highlight a potential advantage of machine learning models in uncovering latent and complex clinical risk factors.

The exploration of nonlinear effects and interaction effects among feature variables in this study revealed the complexity of EUGR risk mechanisms, aspects that are difficult to address with traditional logistic regression models. Currently, literature reporting such nonlinear and interactive relationships in risk prediction studies for EUGR in preterm infants is scarce. Our analysis using SHAP dependence plots and interaction values indicated a clear nonlinear association between birth weight and EUGR risk, while also uncovering significant interaction effects, such as those between birth weight and gestational age, and between breastfeeding and PDA. These findings have theoretical implications. On one hand, they provide data-driven confirmation of the complex synergistic and antagonistic effects among EUGR risk factors. On the other hand, they offer a solid rationale for our choice of the XGBoost model, a tree-ensemble method capable of automatically capturing nonlinearities and interactions [25]. The results of this study validate the correctness of this choice, as the XGBoost model achieved satisfactory predictive performance in the validation set, outperforming the traditional logistic model.

By implementing the XGBoost model into a web-based EUGR risk calculator, this study successfully transitioned the predictive model from theory towards clinical practice. Several risk prediction tools for EUGR have been reported in the perinatal and neonatal fields. Early studies primarily focused on building logistic linear prediction models or providing static nomograms [9, 10]. In recent years, some risk prediction tools based on machine learning have emerged, utilizing methods like random forests or gradient boosting to predict EUGR or other adverse outcomes, often demonstrating model performance through metrics like AUC and DCA, though most remain focused on the model development itself [26, 27]. Some studies have built interpretable prediction models for maternal and infant outcomes using AI or machine learning combined with SHAP but often remain within a research environment, not fully developed into clinically usable tools [28]. This study attempted to operationalize the prediction model further, enabling direct risk estimation by clinicians via a web browser. Through the web calculator, healthcare providers can quickly obtain individualized EUGR risk probabilities simply by entering relevant variables on the webpage, without needing complex formula calculations or chart consultations. The interface also automatically highlights the specific risk factors unique to each predicted individual. The development of this EUGR calculator not only delivers the algorithmic results of a high-performance model but also, through its user-friendly interface and clear interpretability, embeds the model into the clinical workflow. Clinicians can not only obtain a high-risk determination but also, based on the risk factor weights revealed by SHAP analysis, understand the sources of risk, thereby facilitating the development of more targeted nutritional support or clinical monitoring plans. This tool lowers the technical barrier for using machine learning models in clinical settings, potentially aiding its adoption in primary care units.

This study has several limitations. First, although the Boruta algorithm identified the main risk factors included in the model, due to research constraints, many potential influencing factors could not be comprehensively incorporated, such as genetic factors, maternal socioeconomic status, education level, ethnicity, place of residence, and postnatal care factors [29, 30]. Second, the study subjects were primarily drawn from the intensive care units of our hospital, encompassing a relatively high proportion of critically ill patients. Whether the incidence of EUGR in this hospitalized cohort is representative of the local average level requires further verification. As a retrospective study relying on past medical records, potential information bias exists, such as incomplete recording or inconsistent standards for certain variables. The calculator is currently developed based on single-center retrospective data and has not undergone systematic prospective validation across different regions and populations. Its generalizability to primary hospitals or remote follow-up settings remains uncertain.

In conclusion, this study successfully developed and validated an interpretable XGBoost-based model and a corresponding web calculator for the early prediction of EUGR risk in preterm infants. The model demonstrates high predictive performance and good interpretability. This study quantified the contribution of variables within the model, provided visual explanations for individual prediction outcomes, and identified significant interaction effects between low birth weight and gestational age, and between breastfeeding and PDA. This model and its associated calculator can assist healthcare professionals in conveniently and rapidly predicting the risk of EUGR occurrence and, guided by individualized visual explanations, formulating targeted intervention measures, thereby potentially helping to reduce the short- and long-term adverse outcomes in preterm infants. Future research directions should focus on further model refinement, multi-regional, multi-center validation to enhance model robustness and generalizability, incorporation of additional risk factors, in-depth exploration of more interaction effects, and ultimately, the provision of more precise and effective guidance for clinical practice.

## Data Availability

All data produced in the present study are available upon reasonable request to the authors

## Supplementary Information

The supplementary material for this article has been submitted along with the manuscript and can be accessed directly within this article.

Supplementary File 1. Full text of Reference 7 (in Chinese). Supplementary File 2. Executable package of the web-based EUGR risk calculator. Supplementary Figure S1. Performance comparison of the five prediction models. Supplementary Figure S2. Receiver operating characteristic (ROC) curves of the five prediction models. Supplementary Figure S3. SHAP dependence plots for the nine selected features. Supplementary Figure S4. Interaction surface plots for selected feature pairs.

## Acknowledgements

Not applicable.

## Clinical trial number

Not applicable.

## Author contributions

ZX was responsible for study conception and design, methodology development, software programming, formal analysis, model validation, data curation, and writing the original draft. CLY provided critical clinical expertise, assisted in data curation, participated in the interpretation of the results, and contributed to the discussion of the findings. JXZ supervised the research, contributed to the methodological design, and critically reviewed and edited the manuscript for important intellectual content. All authors discussed the results, reviewed the manuscript, and approved the final version. JXZ is responsible for the overall content as the guarantor.

## Funding

The author(s) declare that no financial support was received for the research, authorship, and/or publication of this article.

## Availability of data and materials

All data generated or analysed during this study are included in this published article and its supplementary information files. The datasets used and/or analysed during the current study are available from the corresponding author on reasonable request.

## Declarations

### Ethics approval and consent to participate

The study protocol was approved by the Ethics Committee of the Obstetrics and Gynecology Hospital of Fudan University (Approval No.: 2025-41). The need for informed consent was waived by the committee.

### Consent for publication

Not Applicable.

### Competing interests

The authors declare no conflict of interest.

## Notes

### Competing Interest Statement

The authors have declared no competing interest.

### Funding Statement

This study did not receive any funding

### Author Declarations

Ethics committee/IRB of Obstetrics and Gynecology Hospital, Fudan University gave ethical approval for this work.

