## Supplementary figures and images for "Development and validation of an XGBoost model with SHAP-based interpretability and a web-based calculator for predicting extrauterine growth restriction in preterm infants"

### Supplementary Figure S1

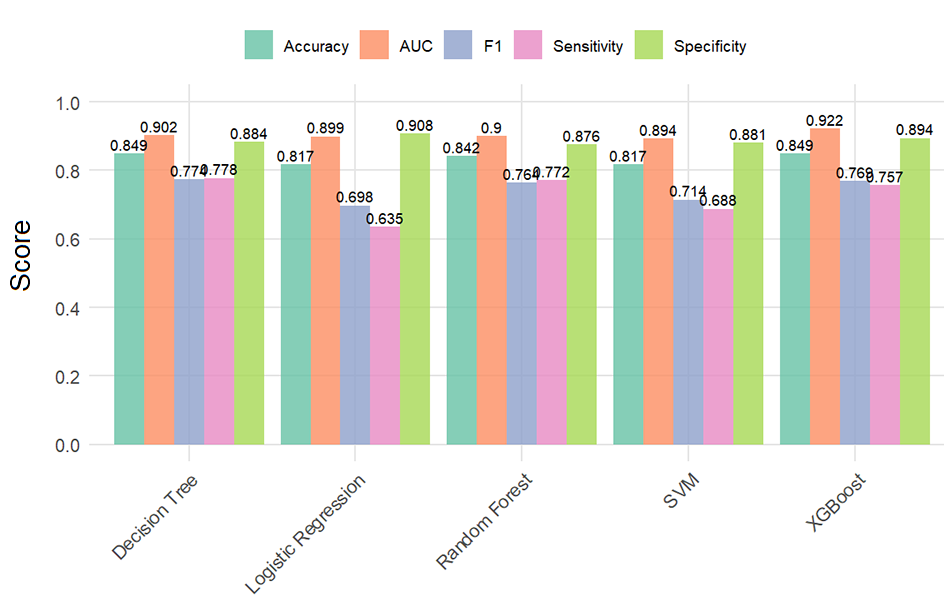

### Supplementary Figure S2

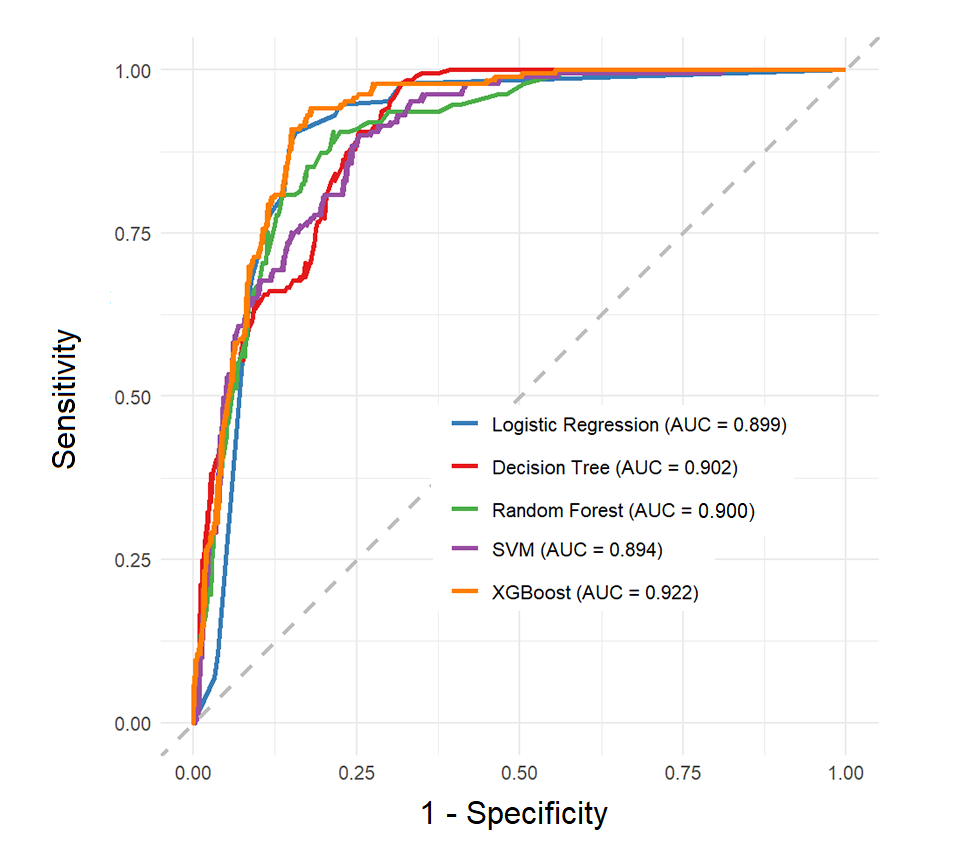

### Supplementary Figure S3

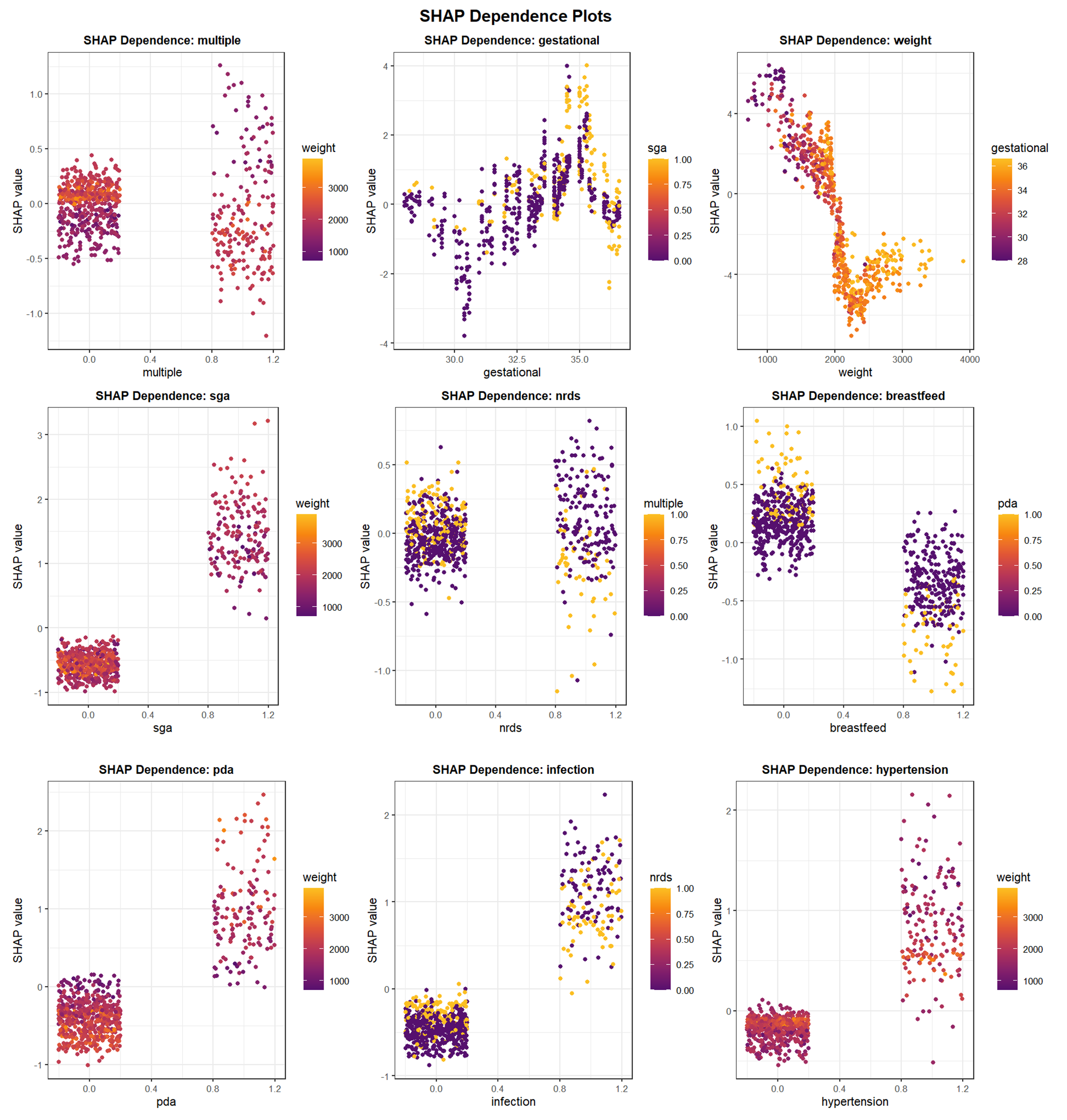

### Supplementary Figure S4

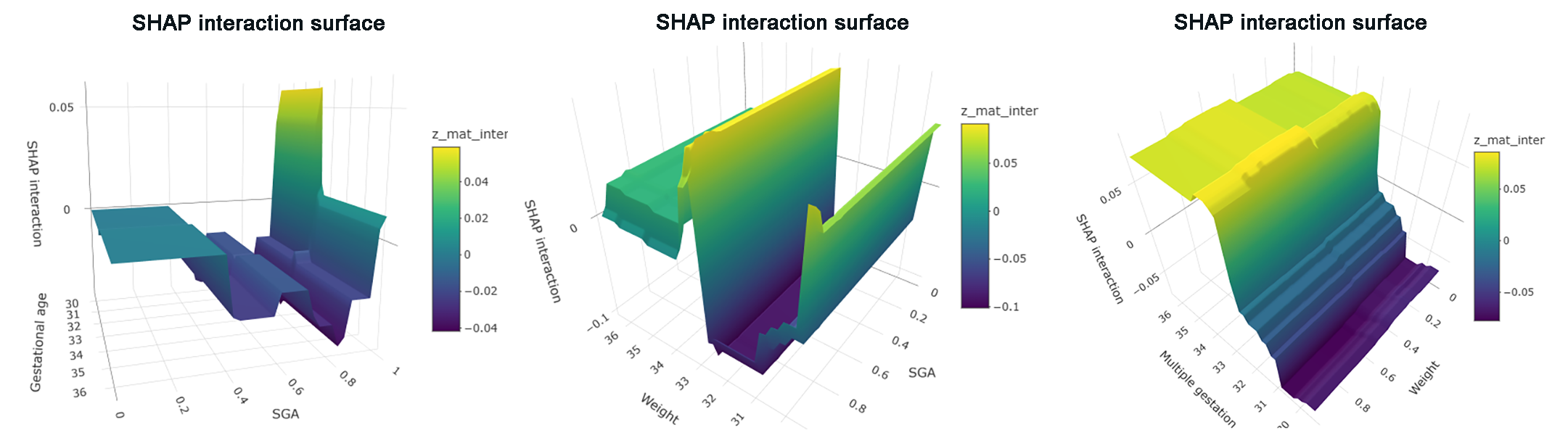
