## Supplementary File 2 for "Development and validation of an XGBoost model with SHAP-based interpretability and a web-based calculator for predicting extrauterine growth restriction in preterm infants": eurg_calculator.py

```
import streamlit as st
import xgboost as xgb
import pandas as pd
import numpy as np
import matplotlib.pyplot as plt
import json
from PIL import Image

# Page configuration
st.set_page_config(
    page_title="EUGR Risk Calculator",
    page_icon="🏥",
    layout="centered",
    initial_sidebar_state="expanded"
)

# App title and description
st.title("🏥 Early Postnatal Growth Restriction (EUGR) Risk Prediction for Preterm Infants")
st.markdown("""
Based on XGBoost Machine Learning Model and SHAP Explainable AI Technology  
**Input early clinical characteristics of preterm infants to assess EUGR risk in real-time**
""")

# Sidebar - Input parameters
st.sidebar.header("📋 Please Enter Clinical Characteristics")

# Create input components based on 9 features
st.sidebar.subheader("Basic Characteristics")

multiplep = st.sidebar.selectbox(
    "Multiple Pregnancy",
    ["No", "Yes"],
    help="Whether it is a multiple pregnancy"
)

gestational = st.sidebar.slider(
    "Gestational Age (weeks)",
    min_value=28.0,
    max_value=36.0,
    value=34.0,
    step=0.1,
    help="Gestational age of preterm infant, range 28-36 weeks"
)

weight = st.sidebar.number_input(
    "Birth Weight (g)",
    min_value=500,
    max_value=4000,
    value=2000,
    step=10,
    help="Birth weight of preterm infant"
)

sga = st.sidebar.selectbox(
    "Small for Gestational Age (SGA)",
    ["No", "Yes"],
    help="Birth weight below the 10th percentile for gestational age"
)

st.sidebar.subheader("Complications and Feeding")

nrds = st.sidebar.selectbox(
    "Neonatal Respiratory Distress Syndrome (NRDS)",
    ["No", "Yes"],
    help="Whether diagnosed with neonatal respiratory distress syndrome"
)

breastfeed = st.sidebar.selectbox(
    "Breastfeeding",
    ["No", "Yes"],
    help="Whether breastfeeding after birth"
)

pda = st.sidebar.selectbox(
    "Patent Ductus Arteriosus (PDA)",
    ["No", "Yes"],
    help="Whether diagnosed with patent ductus arteriosus"
)

infection = st.sidebar.selectbox(
    "Maternal GBS Infection",
    ["No", "Yes"],
    help="Whether mother has Group B Streptococcus infection"
)

hypertension = st.sidebar.selectbox(
    "Maternal Gestational Hypertension",
    ["No", "Yes"],
    help="Whether mother has gestational hypertension"
)


# Convert categorical variables to numerical values
def preprocess_input(multiplep, gestational, weight, sga, nrds, breastfeed, pda, infection, hypertension):
    """Convert user input to numerical format required by the model"""

    # Categorical variable mapping: Yes=1, No=0
    multiplep_encoded = 1 if multiplep == "Yes" else 0
    sga_encoded = 1 if sga == "Yes" else 0
    nrds_encoded = 1 if nrds == "Yes" else 0
    breastfeed_encoded = 1 if breastfeed == "Yes" else 0
    pda_encoded = 1 if pda == "Yes" else 0
    infection_encoded = 1 if infection == "Yes" else 0
    hypertension_encoded = 1 if hypertension == "Yes" else 0

    # Feature order as used during model training
    features = [
        multiplep_encoded,  # multiplep
        gestational,  # gestational
        weight,  # weight
        sga_encoded,  # sga
        nrds_encoded,  # nrds
        breastfeed_encoded,  # breastfeed
        pda_encoded,  # pda
        infection_encoded,  # infection
        hypertension_encoded  # hypertension
    ]

    return features


# Feature names
feature_names = [
    'multiplep', 'gestational', 'weight', 'sga', 'nrds',
    'breastfeed', 'pda', 'infection', 'hypertension'
]

# Prediction button
if st.sidebar.button("🚀 Calculate EUGR Risk", use_container_width=True):

    try:
        # Load model
        model = xgb.Booster()
        model.load_model(r'E:\XGBoost_model.json')  

        # Preprocess input data
        input_features = preprocess_input(
            multiplep, gestational, weight, sga, nrds,
            breastfeed, pda, infection, hypertension
        )

        # Create DataFrame
        input_df = pd.DataFrame([input_features], columns=feature_names)

        # Convert to DMatrix format
        dmatrix = xgb.DMatrix(input_df)

        # Make prediction
        prediction_proba = model.predict(dmatrix)[0]

        # Display main results
        st.header("🎯 Risk Assessment Results")

        # Risk probability display
        col1, col2 = st.columns([1, 2])

        with col1:
            st.metric(
                label="**Predicted EUGR Risk**",
                value=f"{prediction_proba * 100:.1f}%",
                delta=None
            )

        with col2:
            # Risk progress bar
            st.write("**Risk Level:**")
            if prediction_proba < 0.3:
                risk_level = "Low Risk"
                risk_color = "🟢"
                bar_color = "green"
            elif prediction_proba < 0.6:
                risk_level = "Moderate Risk"
                risk_color = "🟡"
                bar_color = "orange"
            else:
                risk_level = "High Risk"
                risk_color = "🔴"
                bar_color = "red"

            st.progress(float(prediction_proba))
            st.write(f"{risk_color} **{risk_level}**")

        # Risk interpretation
        st.subheader("📝 Clinical Interpretation")

        if prediction_proba < 0.3:
            st.success("""
            **Low Risk Indication:**  
            • Current EUGR risk for the infant is low  
            • Continue current feeding and care plan  
            • Regularly monitor growth and development indicators
            """)
        elif prediction_proba < 0.6:
            st.warning("""
            **Moderate Risk Indication:**  
            • Infant has a certain level of EUGR risk  
            • Strengthen nutritional monitoring  
            • Consider optimizing feeding strategies  
            • Closely follow up on weight gain
            """)
        else:
            st.error("""
            **High Risk Indication:**  
            • Infant is at high risk for EUGR, requires special attention  
            • Immediately develop an enhanced nutritional support plan  
            • Actively manage related complications  
            • Strengthen multidisciplinary team collaboration
            """)

        # Feature impact analysis
        st.subheader("🔍 Key Risk Factor Analysis")

        # Simple rule-based analysis based on feature values
        risk_factors = []
        protective_factors = []

        if sga == "Yes":
            risk_factors.append("Small for Gestational Age (SGA)")
        if weight < 1500:
            risk_factors.append(f"Low Birth Weight ({weight}g)")
        if pda == "Yes":
            risk_factors.append("Patent Ductus Arteriosus (PDA)")
        if breastfeed == "Yes":
            protective_factors.append("Breastfeeding")
        if infection == "Yes":
            risk_factors.append("Maternal GBS Infection")
        if hypertension == "Yes":
            risk_factors.append("Maternal Gestational Hypertension")

        col_risk, col_protect = st.columns(2)

        with col_risk:
            if risk_factors:
                st.write("**🔼 Risk-Increasing Factors:**")
                for factor in risk_factors:
                    st.write(f"• {factor}")
            else:
                st.write("**✅ No Significant Risk Factors**")

        with col_protect:
            if protective_factors:
                st.write("**🔽 Protective Factors:**")
                for factor in protective_factors:
                    st.write(f"• {factor}")
            else:
                st.write("**ℹ️ No Significant Protective Factors**")

        # Clinical recommendations
        st.subheader("💡 Personalized Clinical Recommendations")

        suggestions = []
        if weight < 1500:
            suggestions.append("• Initiate **enhanced nutritional support** protocol, closely monitor daily weight gain")
        if pda == "Yes":
            suggestions.append("• Recommend cardiology consultation to evaluate **PDA management** necessity")
        if breastfeed == "No":
            suggestions.append("• Actively encourage and support **breastfeeding**, consider colostrum oral care")
        if sga == "Yes":
            suggestions.append("• Develop **individualized catch-up growth** nutrition plan")

        if suggestions:
            for suggestion in suggestions:
                st.write(suggestion)
        else:
            st.info("• Continue current care protocol, regularly monitor growth and development indicators")

    except Exception as e:
        st.error(f"Error during calculation: {str(e)}")
        st.info("Please ensure the model file 'XGBoost_model.json' is in the correct path")

else:
    # Initial state display
    st.info("💡 Please enter clinical characteristics on the left, then click 'Calculate EUGR Risk'")

    # Display feature description
    with st.expander("📖 Clinical Feature Description"):
        st.markdown("""
        **Feature Definitions:**
        - **Multiple Pregnancy**: Current pregnancy is twins or multiples
        - **Gestational Age**: Gestational age at birth of preterm infant (28-36 weeks)
        - **Birth Weight**: Birth weight of preterm infant (grams)
        - **Small for Gestational Age (SGA)**: Birth weight below the 10th percentile for gestational age
        - **Neonatal Respiratory Distress Syndrome (NRDS)**: Diagnosis of neonatal respiratory distress syndrome
        - **Breastfeeding**: Breastfeeding after birth
        - **Patent Ductus Arteriosus (PDA)**: Diagnosis of patent ductus arteriosus
        - **Maternal GBS Infection**: Mother with Group B Streptococcus infection
        - **Maternal Gestational Hypertension**: Mother with gestational hypertension
        """)

# Footer
st.markdown("---")
st.markdown(
    "<div style='text-align: center; color: gray;'>"
    "Based on XGBoost Machine Learning Model • For Clinical Reference Only"
    "</div>",
    unsafe_allow_html=True
)
```
